# Identification of fibroinflammatory and fibrotic transcriptomic subsets of human sclerotic cutaneous chronic graft-versus-host disease

**DOI:** 10.1101/2022.12.07.22283119

**Authors:** Rachel K. Rosenstein, Jeremy J. Rose, Stephen R. Brooks, Wanxia L. Tsai, Massimo Gadina, Steven Z. Pavletic, Keisuke Nagao, Edward W. Cowen

## Abstract

Cutaneous sclerotic chronic graft-versus-host disease (cGVHD) is a common and highly morbid complication of allogeneic hematopoietic stem cell transplantation (HSCT). We identified genes that are significantly upregulated in the skin of sclerotic cGVHD patients (n = 17) compared to HSCT patients without sclerotic cGVHD (n = 9) by bulk RNA sequencing. This revealed two transcriptomic groups of affected patients: those with fibrotic and inflammatory/Th1 gene expression, the fibroinflammatory group, and those with predominantly fibrotic/TGFβ-associated expression, the fibrotic group. This transcriptomic heterogeneity was also associated with histopathologic heterogeneity, with the fibroinflammatory sub-cluster showing more histopathologic features of epidermal cGVHD and inflammation. Further study will help elucidate if these gene expression and histopathologic findings can be used to tailor treatment decisions. Multiple highly induced genes in the skin (*SFRP4, SERPINE2, COMP*) were also found to be significantly induced in sclerotic cGVHD patient plasma (n = 16) compared to control HSCT patients without sclerotic cGVHD (n = 17), suggesting these TGFβ and Wnt pathway mediators as candidate blood biomarkers of disease.

## TEXT

Cutaneous sclerotic chronic graft-versus-host disease (cGVHD) causes significant morbidity following allogeneic hematopoietic stem cell transplantation (HSCT) (Inamoto et al., 2013). A proposed model for the initiation and progression of cGVHD begins with tissue injury and innate immune activation, evolving into alloreactive dysregulated adaptive immune responses, and, ultimately, inappropriate tissue repair responses, resulting in fibrosis (Zeiser and Blazar, 2017). Management is complicated by heterogeneous clinical presentations and variable response to immunosuppressive therapy. Specific treatments targeting fibrotic mechanisms and prognostic biomarkers are lacking (Wolff et al., 2021).

In allogeneic HSCT patients with sclerotic cGVHD (Supplementary Tables S1-S3), we identified genes that are significantly upregulated in the skin of patients (n = 17) compared to control HSCT recipients without sclerotic cGVHD (n=9), by bulk RNA sequencing (Figure 1a,b). Pathway analysis revealed Th1 pathway (induction of e.g. *TBX21, STAT1, IL12RB1, JAK3)*, phagosome formation (*FCGR1A, FCGR3A, CXCL10)*, and neuroinflammation signaling (*MMP9, TLR7, TLR8, TREM2)*, among the most upregulated inflammatory pathways, along with the fibrotic pathways of pulmonary fibrosis (*THBS1, MMP11, MMP1, FN1)*, wound healing (*COL11A1, COL10A1, COL8A1)*, hepatic fibrosis (*SERPINE1, COL3A1, CCN2)*, and tumor microenvironment *(TGFB1, TGFB3, TNC)*, and lower induction of Th2 pathway *(IL4R, TNFSF4)* (Figure 1c,d). Upstream regulator analysis suggested activation of inflammatory cytokines (TNF, OSM, IL1β, IFNα), Th1 mediators (IFNγ, STAT1), and TGFβ1 (Figure 1e). Overall, these data support a role for T cells and macrophages in the pathogenesis of cGVHD. Whereas Th17 signaling has been implicated in cGVHD (Radojcic et al., 2010; Brüggen et al., 2014; MacDonald et al., 2017), and Th2 signaling has been most associated with diffuse systemic sclerosis (dSSc) (Hasegawa et al., 1997; Greenblatt et al., 2012; Shah et al., 2022), we found that Th1 signaling was predominant in human sclerotic cGVHD, as also reported by others (Zouali et al., 2022).

**Figure 1:**
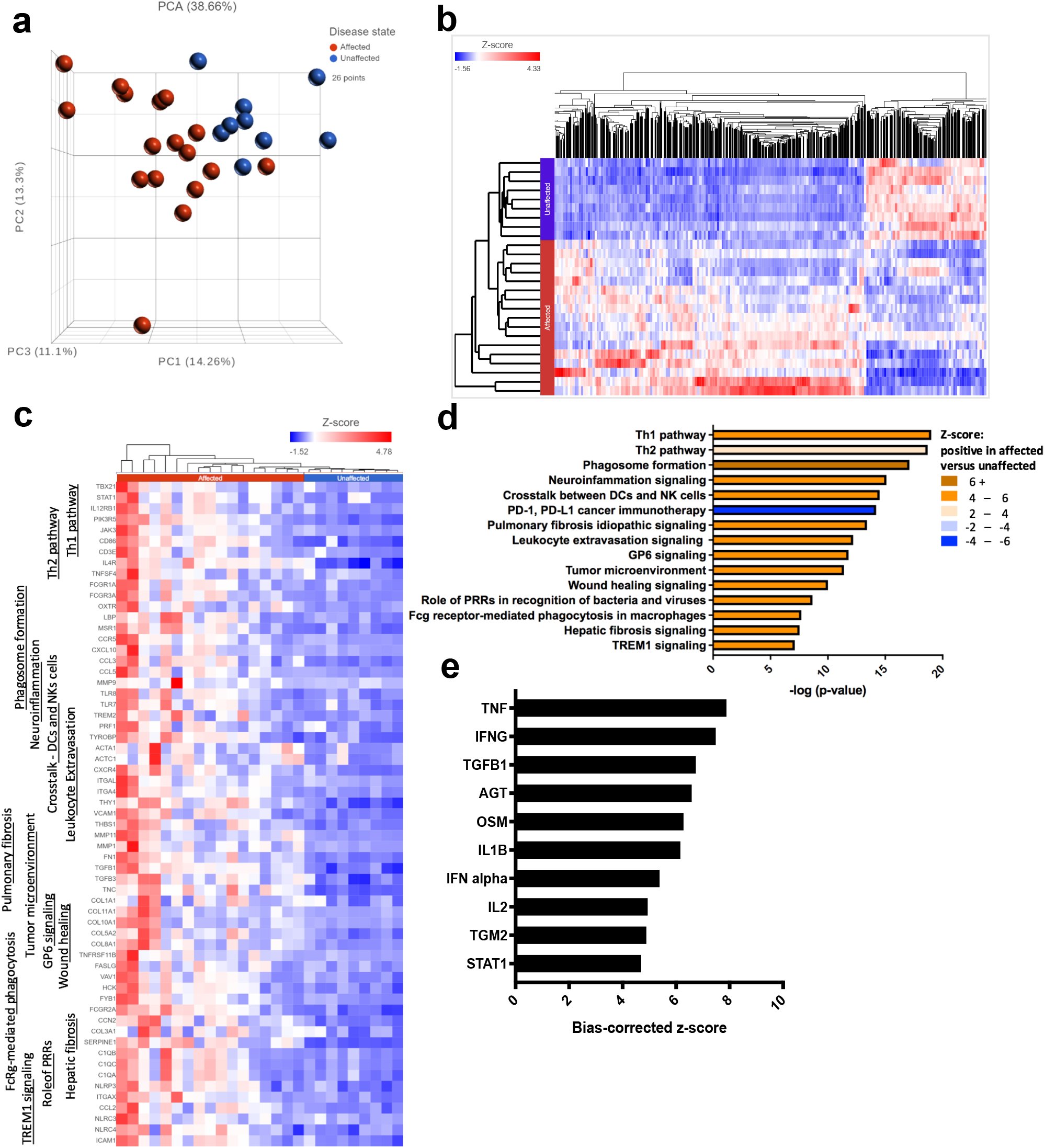
Th1, phagocytic, and fibrotic pathway genes are induced in sclerotic cGVHD. **skin**. (a) PCA plot displaying separation of affected (HSCT patients with sclerotic cGVHD) and unaffected (HSCT patients without sclerotic cGVHD) samples. (b) Heatmap displaying the most differentially expressed genes between affected and unaffected samples, those with a fold change < -1.5 or > 1.5 and FDR step up ≤0.05 resulting in 1173 upregulated and 483 downregulated genes. (c) Heatmap illustrating the gene expression associated with many of the most significantly activated canonical pathways in affected skin. (d) Activated canonical pathways in affected skin. (e) Predicted upstream regulators in affected skin. DCs, dendritic cells; NK, natural killer; PRR, pattern recognition receptor.

While PCA analysis revealed clear separation of unaffected and affected samples, two separate sub-clusters of sclerotic cGVHD were evident (Figures 1a and 2a). The upper affected group was enriched in samples from patients with histopathologic evidence of both sclerotic and epidermal cGVHD, including necrotic keratinocytes and significant inflammation, and a history of stable or progressive disease, while all of the samples without significant inflammation were in the lower affected group (Figure 2b). The highest number of differentially expressed genes was found between the upper affected group and the unaffected group (blue) (e.g. *CXCL9-11,13, GZM*s) (Figure 2c). The intersection of the upper affected vs unaffected groups (blue) and the upper vs lower affected groups (green) had the greatest number of genes (318) (*CXCR3, JAK3, PRF1)*, suggesting that the lower affected and unaffected groups are most similar. Several of the most significantly upregulated genes from the aggregate analysis were identified in both the upper vs unaffected and lower vs unaffected (pink) comparisons (*SERPINE1, SERPINE2, COMP, SFRP4, SFRP2)*, suggesting these are core genes that are uniformly induced in sclerotic cGVHD. Upstream regulator and canonical pathway analysis revealed differences among the groups: the upper vs lower and upper vs unaffected pathway analysis was predominantly inflammatory, while the lower vs unaffected comparison was primarily fibrotic (Figure 2d,e), suggesting that these transcriptomic clusters may represent different functional groups that characterize clinically relevant subsets.

**Figure 2:**
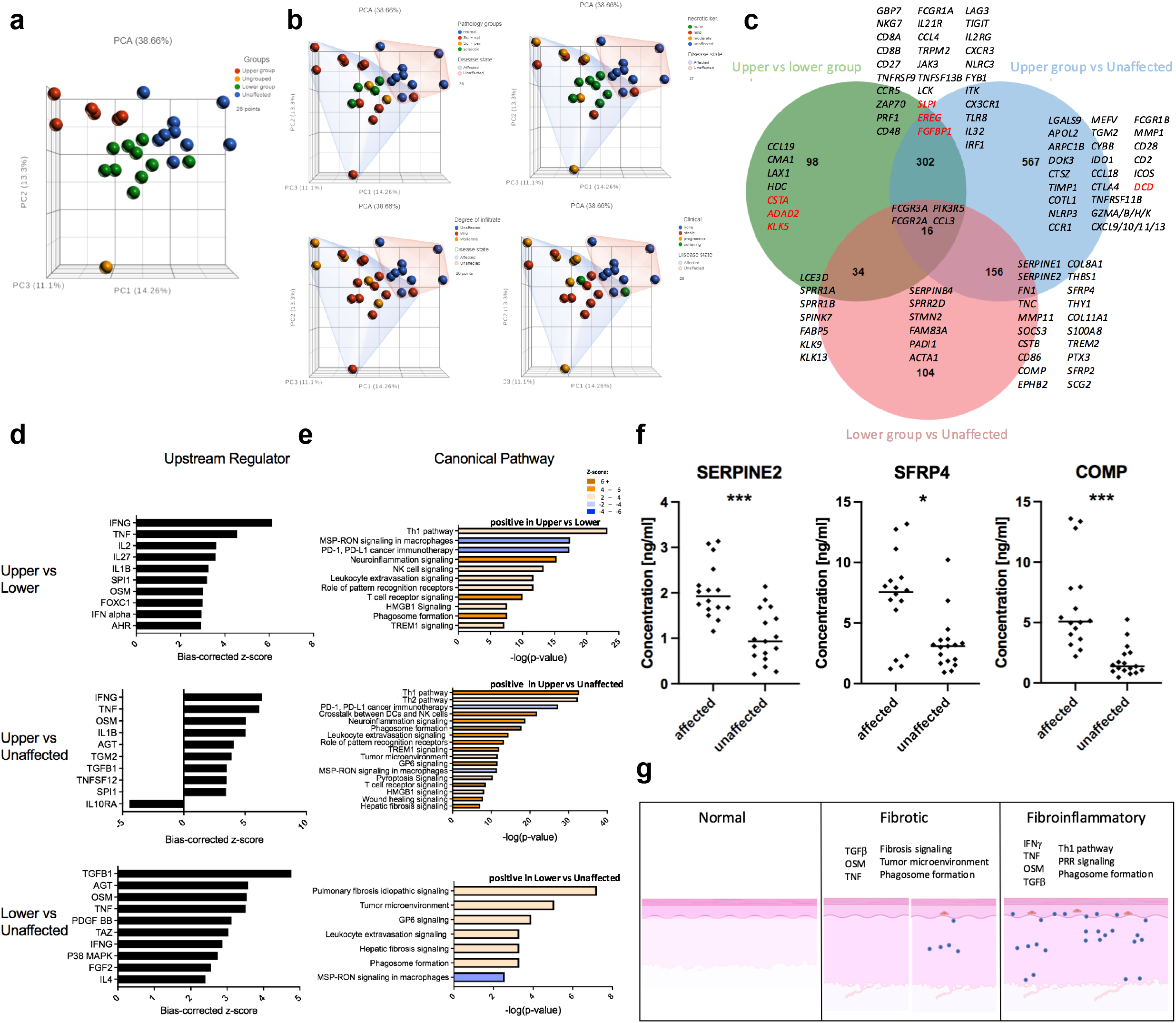
Sclerotic cGVHD skin samples can be divided into fibroinflammatory and fibrotic subsets, while fibrotic genes are uniformly induced in skin of cGVHD patients with fibrosis, and significantly induced in the plasma of sclerotic cGVHD patients. (a) PCA plot displaying two clusters of cGVHD specimens. (b) PCA plots displaying correlation between clusters, clinical course and histopathology. (c) Venn diagram displaying overlapping and non-overlapping genes induced (black) and repressed (red) between the groups: upper vs lower, upper vs unaffected and lower vs unaffected. (d) Predicted upstream regulators. (e) Associated canonical pathways. (f) Plasma levels (ng/ml) of SERPINE2, SFRP4, and COMP in sclerotic cGVHD patients and no cutaneous cGVHD HSCT controls. * = P < 0.01 and *** = P < 0.0001. (g) Schematic of signaling and histopathology. Fibrotic and fibroinflammatory groups have thickened collagen, but fibrotic specimens may demonstrate minimal inflammation (lymphocytes-blue cells) and no epidermal changes (ex. necrotic keratinocytes-red cells) compared to fibroinflammatory specimens, which have inflammation and/or epidermal cGVHD findings.

Multiple fibrosis and TGFβ-associated genes were uniformly induced in affected samples (Supplementary Figure S1a), while inflammatory genes (Th1, CD8-associated) were highly induced, but less uniformly, with an apparent association with the density of the inflammatory infiltrate (Supplementary Figures S1b,c). There was also differential expression of regulatory molecules of Wnt signaling (Supplementary Figure S1d), which have also been used to define different human fibroblast populations (Tabib et al., 2018). *SFRP4*, a Wnt regulator, was highly induced in sclerotic cGVHD skin, was previously found to be induced in diffuse systemic sclerosis (dSSc) skin (Bayle et al., 2008), and is a marker of scleroderma-associated myofibroblasts (Tabib et al., 2021). Overall, higher levels of *SFRP4* were expressed in samples with inflammatory changes compared to those with sclerotic-only findings (Supplementary Figure S2).

Of note, overall changes in gene expression did not correlate with body surface area involvement (Supplementary Figure S3a). While there appeared to be separation between samples from patients recently diagnosed with sclerosis (in the prior year, ‘early’) compared to greater than a year (‘late’), this study was not powered to identify significant differences (Supplementary Figure S3b).

Proteins encoded by highly-induced genes in the skin (*SFRP4, SERPINE2, COMP*) were also highly induced in the plasma of sclerotic cGVHD patients (n = 16) compared to control HSCT patients without sclerotic cGVHD (n = 17) (Figure 2f). Expression of COMP (cartilage oligomeric matrix protein), a member of the PI3K-Akt pathway, has been shown to correlate with the skin fibrosis score in dSSc patients (Moon et al., 2019). SERPINE2 (glia-derived nexin) is a TGFβ-regulated serine protease inhibitor, induced in multiple types of visceral fibrosis (Li et al., 2016). Studies should evaluate the performance of these candidates, individually or in aggregate, as pharmacodynamic or prognostic biomarkers of sclerotic cGVHD.

Overall, this study reveals two transcriptomic groups of sclerotic cGVHD: fibroinflammatory, with fibrotic and inflammatory/Th1 gene expression, and fibrotic, with predominantly fibrotic/TGFβ-associated expression (Figure 2g). This transcriptomic heterogeneity is also associated with histopathologic heterogeneity, with the fibroinflammatory group showing more features of epidermal cGVHD. Additionally, these patients were more likely to report stable or progressive disease than improvement. All patients without inflammation on histopathology were contained in the fibrotic subset. Further study will help elucidate if these gene expression and histopathologic findings can be used to tailor treatments. Potentially, patients with the fibroinflammatory phenotype would be more likely to respond to immunosuppressive medications, whereas these agents may have less utility for patients with the fibrotic phenotype, who might benefit from earlier treatment with a Rho-associated coiled-coil-containing protein kinase 2 (ROCK2) inhibitor, which has anti-fibrotic and anti-inflammatory functions (Zhou et al., 2013). Correlation between gene expression, histopathology, and drug response will be necessary to personalize treatments and optimize clinical care for patients with this highly morbid cGVHD manifestation (Wolff et al., 2021).

## Data Availability

All data produced in the present study are available upon reasonable request to the authors

https://www.ncbi.nlm.nih.gov/geo/query/acc.cgi?acc=GSE216645

## ABBREVIATIONS

(cGVHD): chronic graft-versus-host disease
(HSCT): allogeneic hematopoietic stem cell transplantation
(PCA): principal component analysis
(dSSc): diffuse systemic sclerosis
(ROCK2): Rho-associated coiled-coil-containing protein kinase 2

## DATA AVAILABILITY STATEMENT

The RNA-seq dataset related to this article will be found at

(GSE216645: https://www.ncbi.nlm.nih.gov/geo/query/acc.cgi?acc=GSE216645). There will be no restrictions on data availability.

## ACKNOWLEDGMENTS

We sincerely thank the patients for participating in these investigations. There was institutional approval of experiments and written informed patient consent. This work was supported by the intramural programs of the National Institute of Arthritis and Musculoskeletal and Skin Diseases and the National Cancer Institute. We appreciate the technical support of the NIAMS Genomic Technology Section and Translational Immunology Section. This work utilized the computational resources of the NIH HPC Biowulf cluster (http://hpc.nih.gov). The Figure 2g schematic was created with Biorender.com.

## AUTHOR CONTRIBUTIONS

Conceptualization: RKR, KCN, EWC; Data analysis: RKR, SRB, WLT; Funding acquisition: SZP, EWC; Investigation: RKR, WLT; Methodology: RKR; Data curation: RKR, JJR; Project administration: RKR, EWC; Resources: JJR, MG, SZP; Supervision: MG, KCN, EWC; Visualization: RKR; Writing-original draft: RKR, EWC; Writing-review and editing: RKR, JJR, SRB, WLT, MG, SZP, KCN, EWC.

