## Supplemental Table 1 for "Identification of fibroinflammatory and fibrotic transcriptomic subsets of human sclerotic cutaneous chronic graft-versus-host disease"

| | | Unaffected (N=9) | Affected (N=17) |
|---|---|---|---|
| **Characteristic** | | | |
| **Median age - years** | | 59 | 52 |
| **Female sex - no. (%)** | | 2 (22) | 8 (47) |
| **Underlying diagnosis no. (%)** | | | |
| | MDS/AML | 2 (22) | 6 (35) |
| | CML | 1 (11) | 0 |
| | ALL | 0 | 3 (18) |
| | CLL | 0 | 1(6) |
| | NHL | 3 (33) | 1 (6) |
| | HL | 0 | 2 (12) |
| | MGUS/MM | 1(11) | 2 (12) |
| | myelofibrosis | 1 (11) | 2 (12) |
| | sickle cell | 1 (11) | 0 |
| **Sex-matched transplant - no. (%)** | | 4 (44) | 5 (29) |
| **HLA-matched transplant - no. (%)** | | 8 (89) | 9 (100) |
| **Time since transplant - (years)** | | 6 | 3.6 |
| **Time since sclerosis onset - (months)** | | N/A | 12-18 |
| | late no. (%) | N/A | 8 (47) |
| **History of acute skin GVHD - no. (%)** | | 7 (78) | 7 (41) |
| **Biopsy site** | arm | 3 (33) | 3 (18) |
| | abdomen | 5 (56) | 9 (53) |
| | back | 1 (11) | 2 (12) |
| | chest | 0 | 1 (6) |
| | thigh | 0 | 2 (12) |
| **Clinical impression of skin** | Stable | N/A | 9 (53) |
| | Worsening | | 6 (35) |
| | Softening | | 2 (12) |
| **Histology** | Mild inflammation | N/A | 12 (71) |
| | Moderate inflammation | N/A | 5 (29) |
| | Mild necrotic ker. | N/A | 5 (29) |
| | Moderate necrotic ker. | N/A | 5 (29) |
| **BSA** | | N/A | 42% |
| **Other GVHD organ involvement** | lung | 4 (44) | 12 (71) |
| | liver | 2 (22) | 11 (65) |
| | ocular | 5(56) | 15 (88) |
| | oral | 5 (56) | 11 (65) |
| | genital | 1 (11) | 5 (29) |
| | joint | 1 (11) | 16 (94) |
| | gastrointestinal | 2 (22) | 4 (24) |
| **Current systemic therapy** | none | 4 (44) | 1 (6) |
| | prednisone | 5 (56) | 13 (76) |
| | ECP | 2 (22) | 8 (47) |
| | tacrolimus/cyclosporine | 1 (11) | 5 (29) |
| | sirolimus | 0 | 6 (35) |
| | etanercept | 0 | 1 (6) |
| | mycophenolate mofetil | 0 | 6 (35) |
| | IVIG | 0 | 1 (6) |

**Supplementary Table S1: Demographic and clinical characteristics summary.** BSA, Body surface area; MDS, myelodysplastic syndrome; AML, acute myeloid leukemia; CML, chronic myeloid leukemia; NHL, non-Hodgkin's lymphoma; HL, Hodgkin's lymphoma; MGUS, monoclonal gammopathy of undetermined significance; MM, multiple myeloma; ker, keratinocyte; ECP, extracorporeal photopheresis; IVIG, intravenous immunoglobulin.
