## Supplemental Table 2 for "Identification of fibroinflammatory and fibrotic transcriptomic subsets of human sclerotic cutaneous chronic graft-versus-host disease"

| Pt | Age range | Sex | Oncologic diagnosis | Transplant type | Time since tx[a] | Time since sclerosis[b] | Acute skin GVHD | Biopsy site | Clinical | Histopathology | Sclerotic BSA | Skin score | Other cGVHD | Current systemic therapy | Prior systemic therapy |
|---|---|---|---|---|---|---|---|---|---|---|---|---|---|---|---|
| U1 | 30-39 | M | AML | sex-m, MUD | 4 | none | yes | upper arm | unaffected | normal | 0 | 0 | lung, liver, ocular, oral | pred | mtx, tacro, ATG, siro, mmf |
| U2 | 50-59 | F | myelofibrosis | sex-m, HLA-m | 2 | none | yes | abdomen | unaffected | normal | 0 | 0 | joint, genital, lung, ocular, oral | pred | tacro |
| U3 | 50-59 | F | T cell lymphoma | partial MUD | 9.5 | none | yes | abdomen | unaffected | normal | 0 | 0 | none | pred | tacro, mmf |
| U4 | 60-69 | M | MDS | sex-mm, HLA-m | 8 | none | yes | arm | unaffected | normal | 0 | 0 | ocular, oral, renal | pred, ECP | mmf, ritux, IVIG |
| U5 | 60-69 | M | non-Hodgkin's lymphoma | sex-mm, partial HLA | 4 | none | unk | flank | unaffected | normal | 0 | 0 | none | none | none |
| U6 | 60-69 | M | mantle cell lymphoma | sex-m, HLA-m | 9 | none | no | arm | unaffected | normal | 0 | 0 | none | none | pred, cyclo, siro |
| U7 | 50-59 | M | CML | sex-mm, HLA-m | 2 | none | yes | abdomen | unaffected | normal | 0 | 0 | lung, liver, GI, ocular, oral | pred, tacro, ECP | cyclo, mmf |
| U8 | 20-29 | M | sickle cell | sex-m, HLA-m | 9 | none | yes | back | unaffected | normal | 0 | 1 | lung, GI, ocular, oral | none | tacro, cyclo, mmf, pred |
| U9 | 60-69 | M | multiple myeloma | sex-mm, HLA-m | 6 | none | yes | abdomen | unaffected | normal | 0 | 0 | none | none | pred, ritux |
| A1 | 50-59 | F | myelofibrosis | sex-m, HLA-m | 5 | >24 | no | arm | subcu sclerosis, stable | mild inflammation, rare dyskeratosis, dermal/subcu sclerosis | 63 | 3 | joint, genital, lung, liver, ocular, oral | pred, tacro, siro, etan, ECP | mmf, ritux |
| A2 | 50-59 | F | MDS/AML | MUD | 2 | 0-6 | no | abdomen | dermal/subcu sclerosis, worsening | moderate inflammation, rare dyskeratosis, dermal/subcu sclerosis | 10 | 2 | joint, genital, lung, liver, ocular, oral | none | pred |
| A3 | 50-59 | M | mastocytosis/MDS | sex-mm, HLA-m | 5 | 12-18 | no | abdomen | dermal sclerosis, possible softening | mild inflammation, rare dyskeratosis, dermal/subcu sclerosis | 41 | 2 | joint, lung, liver, GI, ocular, oral | pred | ECP, ritux, tacro, UVA-1 |
| A4 | 20-29 | M | MDS/AML | sex-mm, HLA-m | 3.5 | 6-12 | no | arm | dermal sclerosis, possible softening | moderate inflammation, dermal and subcu sclerosis | 50 | 3 | joint, ocular | pred, siro, ECP | tacro, photo, etan, cp, mtx |
| A5 | 60-69 | M | CLL | sex-mm, HLA-m | 7 | >24 | no | flank | dermal sclerosis, worsening | mild interface vacuolization, dyskeratosis, dermal sclerosis | 37 | 2 | joint, liver, ocular, oral | pred, ECP | cyclo, mmf, aza |
| A6 | 50-59 | M | AML | sex-m, MUD | 4 | >24 | yes | flank | subcu sclerosis | dermal and subcu sclerosis | 63 | 3 | joint, lung | pred, siro | mmf, tacro, hcq, imatinib, mtx |
| A7 | 30-39 | M | Hodgkin's lymphoma | sex-mm, HLA-m | 3.5 | >24 | yes | flank | subcu sclerosis, possible worsening | mild interface vacuolization, dyskeratosis, dermal sclerosis | 49 | 3 | joint, lung, liver, ocular, oral | mmf | pred, ECP |
| A8 | 30-39 | F | AML | sex-m, HLA-m | 3.5 | 6-12 | no | shoulder/back | dermal sclerosis | dermal sclerosis | 10 | 1 | joint, genital, lung, GI, ocular, oral | pred, cyclo, mmf, ECP | tacro, siro |
| A9 | 30-39 | M | Hodgkin's lymphoma | sex-mm, HLA-m | 3 | 12-18 | no | chest | dermal sclerosis | mild inflammation, dermal sclerosis | 1 | 2 | liver, ocular, oral | pred | tacro, mtx |
| A10 | 50-59 | M | MGUS | sex-mm, HLA-m | 1.5 | 0-12 | no | back | dermal sclerosis | dermal sclerosis | 7 | 3 | joint, liver, ocular, oral | pred, ECP | tacro |
| A11 | 40-49 | F | ALL | MUD | 3 | 0-12 | yes | arm | dermal/subcu sclerosis, progressive | dermal and subcu sclerosis | 66 | 3 | joint, lung, liver, ocular | pred, mmf, ECP | cyclo, ritux |
| A12 | 50-59 | F | multiple myeloma | sex-m, HLA-m | 5 | >24 | yes | abdomen | dermal sclerosis | mild inflammation, dyskeratosis, dermal/subcu sclerosis | 87 | 3 | joint, lung, liver, ocular, oral | pred, mmf, IVIG | tacro, cyclo, etan, ritux, ECP |
| A13 | 50-59 | F | AML | sex-mm, HLA-m | 2 | 6-12 | no | abdomen | dermal sclerosis | interface vacuolozation, rare dyskeratosis, dermal/subcu sclerosis | 74 | 3 | joint, genital, liver, ocular | tacro, siro, ECP | mmf, ritux, pred, imatinib |
| A14 | 20-29 | F | ALL, pre-T cell-type | sex-m, HLA-m | 2 | 0-12 | no | abdomen | dermal sclerosis with scale, stable | mild inflammation, dyskeratosis, dermal sclerosis | 81 | 3 | joint, genital, lung, liver, ocular | pred, siro, mmf | tacro, ritux, ECP |
| A15 | 50-59 | M | myelofibrosis | sex-mm, HLA-m | 4 | >24 | yes | abdomen | dermal/subcu sclerosis, stable | Mild inflammation, interface vacuolization, dermal/subcu sclerosis | 64 | 3 | joint, lung, liver, GI, ocular, oral | pred, siro, ECP | cyclo, mmf, ritux, ECP, DD |
| A16 | 40-49 | M | follicular lymphoma | sex-mm, HLA-m | 4.5 | 6-12 | yes | thigh | dermal sclerosis, recent onset | Moderate interface dermatitis, dyskeratosis, dermal sclerosis | 9 | 3 | joint, lung | tacro, mmf | pred, ritux |
| A17 | 50-59 | F | ALL | sex-mm, HLA-m | 3 | 0-6 | yes | thigh | sclerosis with scale, possibly recent | mild interface vacuolization, rare dyskeratosis, dermal/subcu sclerosis | 5 | 1 | joint, lung, GI, ocular, oral | pred, tacro | ECP |

**Supplementary Table S2: Demographic and clinical characteristics of the patients.**

[a] in years; [b] in months; Pt (patient); sex-mm (sex-mismatched); sex-m (sex-matched); MUD (matched unrelated donor); HLA-m (HLA-matched); unk (unkown); BSA (body surface area); GI (gastrointestinal); pred (prednisone); tacro (tacrolimus); siro (sirolimus); etan (etanercept); mmf (mycophenolate mofetil); cyclo (cyclosporine); mtx (methotrexate); ATG (anti-thymocyte globulin); rituximab (ritux); photo (phototherapy); cp (cyclophosphamide); aza (azathioprine); hcq (hydroxychloroquine); DD (denileukin diftitox).
