## Supplemental Table 3 for "Identification of fibroinflammatory and fibrotic transcriptomic subsets of human sclerotic cutaneous chronic graft-versus-host disease"

| Patient | Disease | Clinical | Time since sclerosis | Pathology group | Degree of infiltrate | Necrotic keratinocytes |
|---|---|---|---|---|---|---|
| U1 | Unaffected | none | none | normal | unaffected | none |
| U2 | Unaffected | none | none | normal | unaffected | none |
| U3 | Unaffected | none | none | normal | unaffected | none |
| U4 | Unaffected | none | none | normal | unaffected | none |
| U5 | Unaffected | none | none | normal | unaffected | none |
| U6 | Unaffected | none | none | normal | unaffected | none |
| U7 | Unaffected | none | none | normal | unaffected | none |
| U8 | Unaffected | none | none | normal | unaffected | none |
| U9 | Unaffected | none | none | normal | unaffected | none |
| A1 | Affected | stable | late | Scl + epi | mild | mild |
| A2 | Affected | progressive | early | Scl + epi | moderate | mild |
| A3 | Affected | softening | late | Scl + epi | mild | mild |
| A4 | Affected | softening | early | Scl + peri | moderate | none |
| A5 | Affected | progressive | late | Scl + epi | mild | moderate |
| A6 | Affected | stable | late | sclerotic | mild | none |
| A7 | Affected | progressive | late | Scl + epi | mild | moderate |
| A8 | Affected | stable | early | sclerotic | mild | none |
| A9 | Affected | stable | late | Scl + peri | mild | none |
| A10 | Affected | stable | early | sclerotic | mild | none |
| A11 | Affected | progressive | early | sclerotic | mild | none |
| A12 | Affected | stable | late | Scl + epi | mild | moderate |
| A13 | Affected | stable | early | Scl + epi | moderate | mild |
| A14 | Affected | stable | early | Scl + epi | mild | moderate |
| A15 | Affected | stable | late | Scl + epi | moderate | none |
| A16 | Affected | progressive | early | Scl + epi | moderate | moderate |
| A17 | Affected | progressive | early | Scl + epi | mild | mild |

**Supplementary Table S3: Clinical and histopathologic correlative characteristics of patients.** U (unaffected); A (affected); Clinical: as determined by patient and physician assessment; early (less than 1 year since sclerosis onset); late (greater than 1 year); sclerotic (sclerotic changes without significant inflammation); Scl + peri (sclerotic changes and perivascular inflammation without significant epidermal changes); Scl + epi (sclerotic changes with necrotic keratinocytes, melanophages, and/or interface dermatitis); normal (normal histopathology); for the degree of infiltrate, mild (scattered immune cells either in a perivascular or interface pattern); moderate (a moderate inflammatory cell infiltrate in the dermis and/or a combination of a perivascular and interface inflammatory cell infiltrate); for necrotic keratinocytes, none (no necrotic keratinocytes identified), mild (at least 1 identified in the pathology section), or moderate (multiple identified in the pathology section).
