## Supplemental Figure 1 for "Identification of fibroinflammatory and fibrotic transcriptomic subsets of human sclerotic cutaneous chronic graft-versus-host disease"

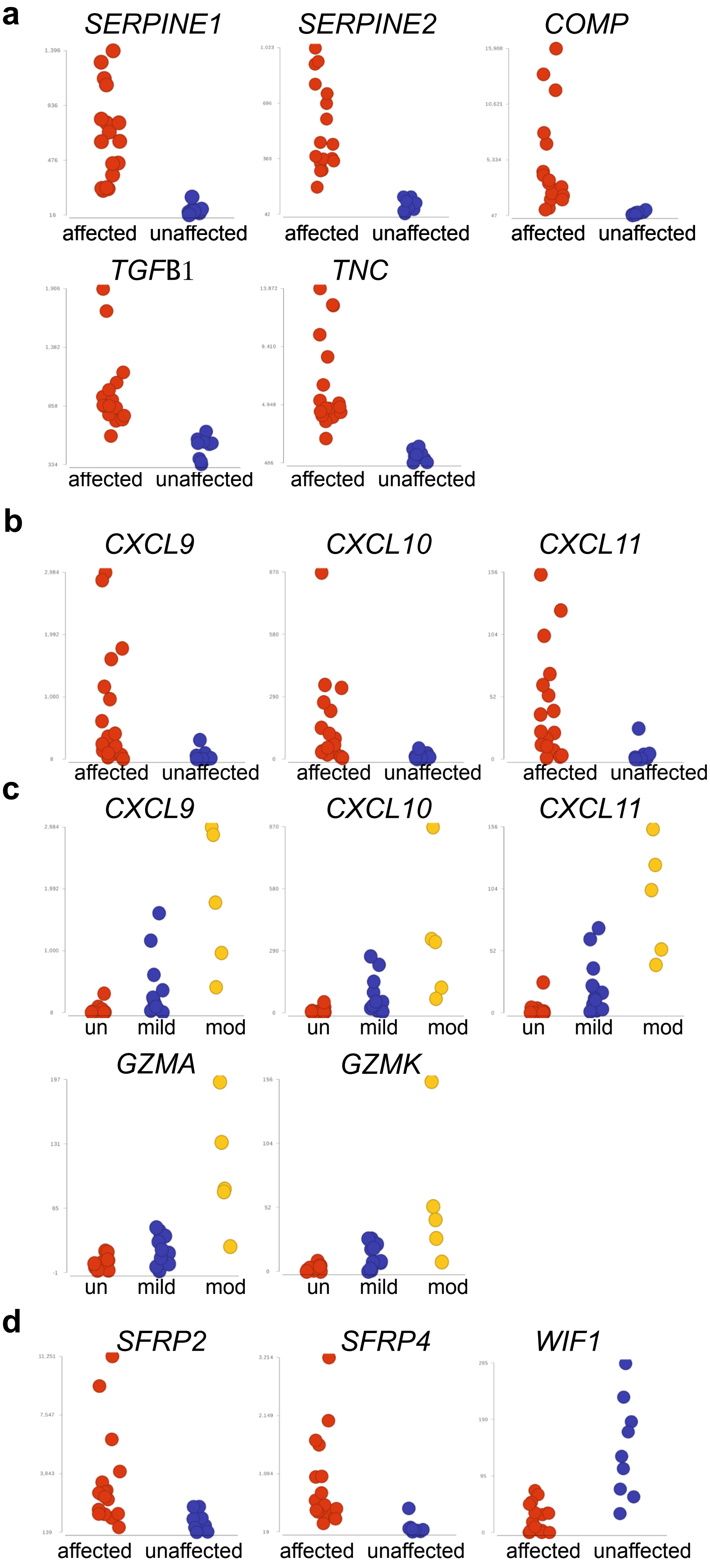


**Supplementary Figure S1: Fibrotic genes are uniformly induced in affected samples while inflammatory gene expression is less uniform and associated with density of inflammation.** (a) Dot plots displaying gene expression of fibrosis and TGFΒ-associated genes in affected skin (HSCT patients with sclerotic cGVHD) and unaffected skin (HSCT patients without sclerotic cGVHD).  (b) Dot plots displaying gene expression of inflammatory chemokine expression in affected and unaffected skin. (c) Dot plots of Th1 and CD8-associated gene expression in affected skin with a mild or moderate inflammatory infiltrate compared to unaffected skin. (d) Dot plots of gene expression of Wnt-pathway associated genes in affected and unaffected skin.
