## Supplemental Figure 2 for "Identification of fibroinflammatory and fibrotic transcriptomic subsets of human sclerotic cutaneous chronic graft-versus-host disease"

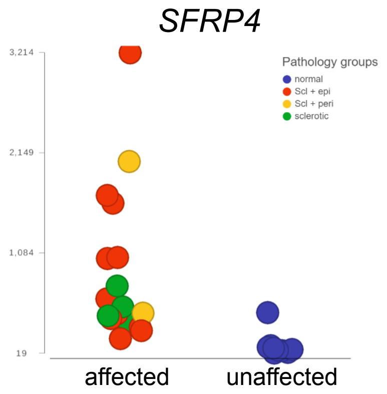


**Supplementary Figure S2: *SFRP4* is more highly induced in samples with inflammatory cells on histopathology compared to those with only sclerosis.** Dot plot of *SFRP4* gene expression in affected skin compared to unaffected skin. Affected specimens were grouped according to the overall histopathology, identifying specimens with changes that were "sclerotic" (sclerotic changes without significant inflammation), "Scl + peri" (sclerotic changes and perivascular inflammation without significant epidermal changes), and "Scl + epi" (sclerotic changes with necrotic keratinocytes, melanophages, and/or interface dermatitis). Unaffected skin was labeled as "normal."
