## Supplemental Figure 3 for "Identification of fibroinflammatory and fibrotic transcriptomic subsets of human sclerotic cutaneous chronic graft-versus-host disease"

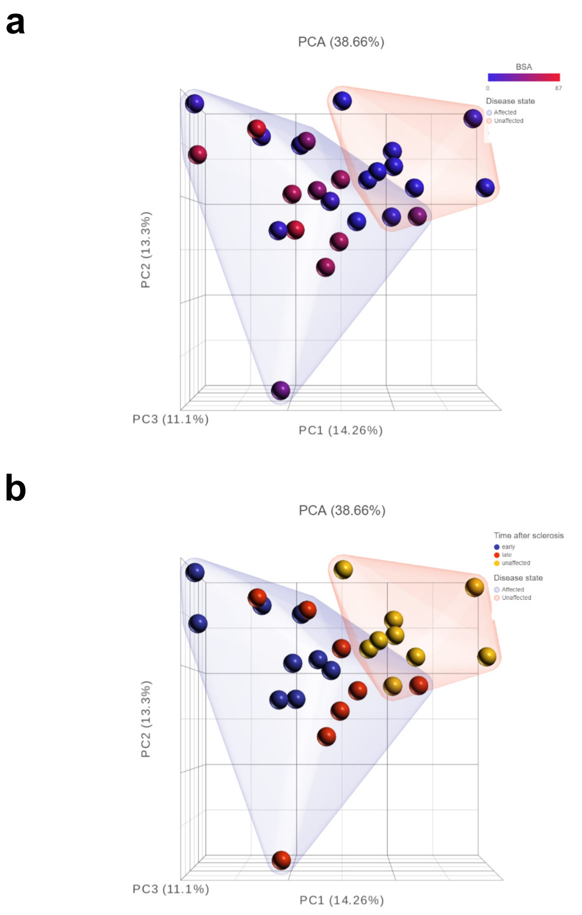


**Supplementary Figure S3: Gene expression does not correlate with body surface area (BSA) involvement while there appears to be a correlation between gene expression and time since onset of sclerosis.** (a) PCA plot displaying affected and unaffected samples, colored according to the BSA.
(b) PCA plot displaying affected and unaffected samples, colored according to the time after sclerosis onset. early (patients diagnosed with sclerotic cGVHD in the prior year); late (patients diagnosed with sclerotic cGVHD over a year before).
