## Supplemental text for "Identification of fibroinflammatory and fibrotic transcriptomic subsets of human sclerotic cutaneous chronic graft-versus-host disease"

**SUPPLEMENTARY MATERIAL**

**Supplementary Table Legends**

**Supplementary Table S1: Demographic and clinical characteristics summary.** BSA, Body surface area; MDS, myelodysplastic syndrome; AML, acute myeloid leukemia;

CML, chronic myeloid leukemia; NHL, non-Hodgkin's lymphoma; HL, Hodgkin's lymphoma;

**Supplementary Materials and Methods**

Human materials

Skin and blood sample collection was performed in accordance with an approved protocol by the Institutional Review Board at the National Cancer Institute (NCT00092235). Individuals provided written informed consent for sample acquisition and subsequent analyses. We have complied with all relevant ethical regulations. Skin biopsies were done on allogeneic HSCT patients with sclerotic cGVHD who were not on high doses of prednisone or targeted cGVHD therapy and compared to normal skin biopsies from allogeneic HSCT patients who were not on high doses of prednisone or targeted cGVHD therapy and had no history of cutaneous cGVHD. For skin biopsies, 6 mm skin punch biopsies were obtained from patients and sections were frozen for RNA sequencing, or placed in formalin for processing for hematoxylin and eosin (H&E) staining by the clinical laboratory. All H&E slides were reviewed to confirm the presence or absence of sclerotic cGVHD by the study clinicians. Additional histopathologic features were categorized. Affected specimens were grouped according to the overall histopathology, identifying specimens with changes that were “sclerotic” (sclerotic changes without significant inflammation), “Scl + peri” (sclerotic changes and perivascular inflammation without significant epidermal changes), and “Scl + epi” (sclerotic changes with necrotic keratinocytes, melanophages, and/or interface dermatitis). Affected specimens were split into those with “mild” inflammation (scattered immune cells either in a perivascular or interface pattern) vs “moderate” inflammation (a moderate inflammatory cell infiltrate in the dermis and/or a combination of a perivascular and interface inflammatory cell infiltrate). Affected specimens were split as determined by the extent of keratinocyte necrosis with “none” (no necrotic keratinocytes identified in the pathology section), “mild” (at least 1 identified in the pathology section), or “moderate” (multiple identified in the pathology section). Clinical features of the patients were also categorized upon chart review. According to patient report and clinician observation of the appearance of lesions, sclerotic skin disease was categorized as “stable,” “progressive,” or “softening.” Two of the nine unaffected patients were female and eight out of the 17 affected patients were female. According to the PCA plots, we did not detect differences among the affected samples as determined by sex.

For the plasma studies, affected patients were allogeneic HSCT patients with sclerotic cGHVD, while unaffected patients were allogeneic HSCT patients who had no history of cutaneous cGVHD, but may have had active extracutaneous organ involvement.

RNA sequencing

Skin biopsies were either flash frozen on dry ice, stored at -80°C, and treated with RNALater-ICE upon use or were treated with RNALater prior to storage at -80°C. Biopsy samples were minced manually and TRIzol solution (Invitrogen) was added to frozen samples, followed by homogenization with PowerGen 125 Homogenizer (Fisher Scientific). After addition of the TRIzol-tissue homogenate to chloroform, samples were shaken, and spun down. The supernatants were collected, placed on ice, mixed with an equal volume of 70% ethanol, and loaded onto RNeasy Mini Spin Columns, followed by completion of the RNeasy Mini (QIAGEN) protocol. Libraries were generated from samples with a RIN number greater than 5.3. 10ng of total RNA was used for the construction of sequencing libraries. Libraries were prepared using the SMARTer Stranded Total RNA-Seq Kit v2-Pico Input Mammalian kit (Takara Bio USA, Inc.) and sequenced on an Illumina HiSeq 3000 (1 x 50bp read length). Libraries were diluted to 3nM and pooled for sequencing. Illumina runs were demultiplexed and converted to FastQ using Casava 1.8.2 and uploaded into Partek Flow.  Raw data was trimmed based on quality score.  Data was mapped to hg38 using STAR 2.7.8a. Features were filtered to exclude features where maximum <=50.  Data were transformed on samples and normalized by Median ratio.  DESeq2 was utilized for differential analysis. Dot plots display median ratio normalized counts.

Canonical pathway and upstream regulator analyses were done using Qiagen Ingenuity Pathway Analysis. For the comparison of unaffected and affected samples, genes with a fold change > 1.5 or < -1.5 and a FDR step up <0.05 were analyzed. For the comparisons of the upper affected group, lower affected group and unaffected samples, genes with a fold change > 2 or < -2 and a FDR step up < 0.01 were analyzed. The venn diagram illustrating the overlap among the differentially expressed genes of these comparisons was produced with jvenn (Bardou et al., 2014).

ELISA (Enzyme Linked Immunosorbent assay)

Human plasma samples were collected and concentrations of COMP/Thrombospondin-5, Secreted Frizzled Related Protein 4 (SFRP4), Glia Derived Nexin/SERPINE2 were measured by ELISA. The assays were performed in duplicate according to the manufacturer’s instructions, using the following kits: for COMP (Catalog # DCMP0, R&D Systems, Inc.), SFRP4 (Product # SEF878Hu, Cloud-Clone Corp.), SERPINE2 (Product # SED381Hu, Cloud-Clone Corp.). Samples were read at wavelengths of 450 and 540nm on a Synergy H1 plate reader (BioTek). Sample concentrations were calculated according to standard curves specific for each analyte. Significance was determined using the Mann-Whitney test.
